# An overview of global monitoring systems for the side effects and adverse events associated with medicinal cannabis use: A scoping review using a systematic approach

**DOI:** 10.1101/2024.02.03.24302171

**Authors:** Rebecca Qi Wang, Yvonne Ann Bonomo, Christine Mary Hallinan

## Abstract

The use of cannabis-based medicines (CBM) as a therapeutic has grown exponentially over the last 5 years in Australia. Prior to this increase, there was significant legislative resistance to the use of CBM for clinical trials, hence pre-clinical data is limited. Safety monitoring systems for CBM are not structured and do not fit easily into the workflow of busy health professionals. Hence, post-marketing surveillance of CBM is patchy. CBM are available in many countries globally and face similar issues in relation to pharmacovigilance. The objective of this review is to answer the following question: What are the systems in place internationally to monitor side effects and adverse events of cannabis use as a medicine?

We used the PICO framework to develop keyword elements, which guided two search queries. Each query contained a different combination of keywords to increase sensitivity and specificity of the search. Both queries were entered into Embase and Scopus for retrieval of quality relevant peer-reviewed literature. Only the second search query, was used for the grey literature. Fifty-four full text articles were included in the review, thirty-nine were from the peer-reviewed search, eight were from the grey literature search, and seven were from citations of relevant texts.

Our search yielded two main forms of monitoring systems: databases and registries, with databases often created by regulatory authorities. There was great variability within these systems, differing in methods of causality assessment, level of detail collected, terminology, and affiliations. Only one monitoring system captured in our search obligated reporting from patients.

VigiBase remains the largest form of centralised monitoring, receiving case reports internationally. Regardless of the scope of VigiBase, there remains heterogeneity of data within the system. As such, our study reaffirms a greater need for a centralised, consistent, and accessible system for the post-marketing surveillance of side effects and adverse events associated with usage of CBM.

**KEY MESSAGES:** *What is already known on this topic:*

- Real-world data is essential for monitoring the side effects and adverse events associated with the use of cannabis-based medicines, given the limited availability of clinical trials, increasing clinical demand, and rising accessibility to unregulated cannabis-based products.
- In some countries, registries and databases exist for post-marketing surveillance of side effects and adverse events at a national level.

*What this review adds:*

- A summary of the current landscape of monitoring systems at an international level, and interactions, and reporting hierarchies that exist between systems.
- An analysis of the content, specificity, and scope of each monitoring system, including an analysis of the reporting type, be it mandatory or spontaneous.

*How this study might affect research, practice, or policy (summarise implications):*

- A robust and standardised system is required for ongoing post-marketing surveillance of the side effects and adverse events associated with usage of cannabis-based medicine.
- Development of a system that is both accessible and well-integrated into healthcare professional clinical workflow is needed.
- Future practice and policy guided by this research can establish a standardised approach for collecting safety data that aligns with the rapid adoption of cannabis-based medicines in clinical settings.

## INTRODUCTION

The emergence of *Cannabis sativa* as a therapeutic can be dated back to 1500 BC, with use becoming widely adopted in the United States by the 19th century (1), (2) .Since then, both recreational and medicinal cannabis have undergone a series of proscription and later decriminalisation processes globally. As of the 21^st^ century, cannabis for medicinal purposes has been legalised in many countries, including the United States, UK, Australia, Canada, Israel, and Netherlands (3).

The *Cannabis* plant contains over 500 different compounds. Of these, 113 are recognised as cannabinoids, where they function as cannabinoid receptors for biological effect (4). Notably, cannabidiol (CBD) and delta-9-tetrahydrocannabinol (THC) are the two main active constituents of over 100 medicinal cannabis products available worldwide. CBM are products containing cannabinoids that are used for a clear therapeutic purpose, rather than recreational purposes. CBM may be obtained on prescription or otherwise, and is generally used for symptomatic control of intractable chronic diseases. These include, but are not restricted to, spasticity in multiple sclerosis (MS), epilepsy, neuropathic pain, cancer-related pain, as well as chemotherapy-induced nausea and vomiting. Emerging evidence is expanding therapeutic usage of CBM to include psychiatric disorders such as anxiety and PTSD, sleep disorders, fibromyalgia, and Parkinson’s disease (2,5-7).

The growing evidence base and media attention has triggered a shift in public paradigms towards acceptance of cannabis as a medicine (8). The increasing community demand for CBMs is apparent in the uptrend of prescription approvals. Over the last 7 years in Australia, there were 949,732 patients who were newly prescribed a specific medicinal cannabis product, biannually via the TGA’s Authorised Prescriber System (9). This uptrend in prescribing rates is further reflected by a percentage increase of 402% in new prescriptions in the six-month period ending January 2022, compared to the six-month period ending January 2023 (Supplementary Material 1) (10).

However, unlike conventional medications, public demand rather than preclinical studies for quality control, have driven increasing clinical uptake (11,12). Given a history of legislative resistance and restrictions in conducting clinical trials with CBMs, gaps remain in the literature surrounding side effects and adverse reactions. Notably, there is limited safety evidence on CBMs for vulnerable populations commonly excluded from clinical trials, such as pregnant women, children, and patients with complex comorbidities (13-15). Additionally, the illicit drug market, over-the-counter availability, and unregulated product commercialisation has created a landscape of products that vary in formulation, strength, route of administration and quality (16-18). As such, growing use necessitates prescriber and consumer vigilance on side effects and adverse reactions.

The discrepancy between available safety evidence, and clinical use warrants rigorous surveillance and process for post-marketing signal detection as surveillance of CBMs. Different countries have undertaken this need in different ways. The objective of this research therefore is to collate an overview of current methods for real-world monitoring of the side effects and adverse events associated with CBM use. Using a systematic search of peer-reviewed databases and grey literature, this review aims to answer the following question: What are the systems in place internationally to monitor side effects and adverse events of cannabis use as a medicine?

## METHODS

### Search Strategy

The PRISMA reporting guidelines were used as a methodological framework to inform the approach to a systematic search of literature (19,20). Five main keyword elements were identified using the PICO framework, and subsequently used to guide development of search terms and inclusion/exclusion criteria (Supplementary Material 2).

Two separate categories of searches were conducted, each with search terms from a different combination of keyword elements (Supplementary Material 3). For the first category, the search query combined terms relating to elements of medical usage, cannabis, monitoring systems, and side effects or adverse events. The second category included cannabis related terms, as well as terms relating to pharmacovigilance, monitoring systems, and medical usage. The first category aimed to increase the specificity of our search, whereas the second category focused on search sensitivity, incorporating the more loosely defined concept of pharmacovigilance, without specific mention of side effects and adverse events. Both categories were used to create searches on 23^rd^ of June 2023, identical across Scopus and Ovid Embase for peer-reviewed publications, with added MESH terms in the latter (Supplementary Material 4).

Category 2 search terms were further used for a grey literature search to supplement our literature database search and maximise the scope of our results. The grey literature search comprised of extracting the first 1000 titles of a Google Scholar search using Category 2 search terms. An identical search for grey literature was applied to Mednar, a medically focussed search engine, to include deep web searches that were not indexed by standard search engines (21-23). Search terms across different keyword elements were combined with Boolean operator “AND” while terms within a keyword element were combined with the Boolean operator “OR”.

Supplementary articles were identified in the references of retrieved papers. All searches were limited to papers published between January 2015 and June 2023; the period in which cannabis legalisation occurred in multiple countries worldwide, triggering the need for widespread monitoring systems (24-26). The search was manually filtered to papers published in the English language, to yield the results shown in the PRISMA flowchart (Figure 1).

All search results from search queries were uploaded into an excel spreadsheet for duplicate removal. Title and abstract screening were performed, subject to inclusion and exclusion criteria. Records were excluded primarily based on relevance and the a priori decision to exclude records with no mention of pharmacovigilance nor a monitoring system in the title and abstract. Secondary google searches were performed for primary sources such as reporting forms, where more specific information on databases were required. Small scale surveys were not considered a formal monitoring system and subsequently excluded. All full text articles identified for inclusion following the screening process were evaluated independently by at least two reviewers. Any points of contention were discussed in meetings and subsequently resolved for a full list of titles for data extraction. RW reviewed all titles, and YB provided a second review. There was discrepancy between the reviewer assessments in <5% of articles, which were subsequently resolved amongst the reviewers, thus not requiring a third reviewer. Any points of contention were discussed in meetings and subsequently resolved for a full list of titles for data extraction.

The PRISMA flowchart (Figure 1) outlines the full search strategy and results.

## RESULTS

Of the 3939 records identified in the initial peer-reviewed database search, 1004 duplicates were removed, with an additional 2889 records excluded following title and abstract screening. The screening process yielded 46 potentially relevant full texts from the peer-reviewed database search, with 3 unable to retrieved, leaving 43 full texts for inclusion. The grey-literature search identified 1127 records. Following duplicate removal and title and abstract screening, 12 full texts were identified. Of these, 1 record was unable to be retrieved. Subsequently, 11 additional records from the grey literature search were identified for full text review, leaving 55 records for full text review. Of the 54 full texts assessed for eligibility, 5 were excluded on the basis of relevance, and 2 were excluded for recreational cannabis use as the study population. 7 additional papers were identified through citations, yielding a total of 54 included records (Supplementary Material 5).

Monitoring systems identified by our search were either registries or databases. Although used interchangeably, there are several distinguishing characteristics between the two (Supplementary Material 6). There was a total of 7 regulatory authority databases and 17 registries captured within our search (Table 1). Of these registries, 8 were smaller registries briefly mentioned in articles, without readily available data and were not analysed in depth (Table 1).

**Table 1.** Monitoring systems captured by search with readily available data for analysis*.

| Databases | Registries |
| --- | --- |
| Italian Phytovigilance Database (27,28) | German Pain e-registry (29-33) |
| FDA Adverse Event Reporting System (FAERS) (28,34-37) | Quebec Cannabis registry (28,38-41) |
| WHO VigiBase (28,42,43) | The Registry (32,44,45) |
| Canada Vigilance Adverse Reaction Online Database (CVAR) (28,37-39,41,46) | Australian Emyria Clinical Registry (47) |
| Eudravigilance European Database of Suspected Adverse Drug Reaction Reports (EDSADR) (48) | UK Medicinal Cannabis Registry (UKMCR) (47,49-67) |
| Drug Commission of the German Medical Association database (DCGMA) (68) | Toxic registry (69-72) |
| TGA Database of Adverse Event Notifications (DAEN) (47,73) | SwissCanOn (74) |
| *CB2 Insights' Clinical network database (75)* | Italian Medicines Agency (AIFA) Registry (32,76,77)** |
| *NotiFACEDRA database (42)* | Project TwentyOne (28,51,67,78,79) |
| *DATAcANN: Database for Cannabinoid Consumption and Study (28)* | *Israeli Multi-Center Registry of Medical Cannabis for Chronic Pain (80)* |
|  | *Spanish Prospective Registry (45)* |
|  | *Minnesota Department of Health: Medicinal Cannabis Registry (28)* |
|  | *Children's Hospital of Philadelphia: Medical Cannabis Registry (28)* |
|  | *Canadian Paediatric Surveillance Program (28)* |

\*Monitoring systems in italics were systems captured by search that did not have readily available data for analysis.
\*\*Known as a registry but governed by a regulatory authority (Italian Medicines Agency)

Notably, many registries input data into larger databases. Interactions between registries and databases are captured in Supplementary Material 7 (42).

### Monitoring System Characteristics

#### 1. Primary purpose

There were two distinct purposes for establishment of monitoring systems. Some systems were created as broader forms of post-marketing surveillance to inform safety and regulation. These include all aforementioned databases and all registries with the exception of five. These registries were created as data collection for observational studies, with post-marketing surveillance as a secondary aim from the research. As such, they are categorised as research registries (Table 2).

**Table 2.**
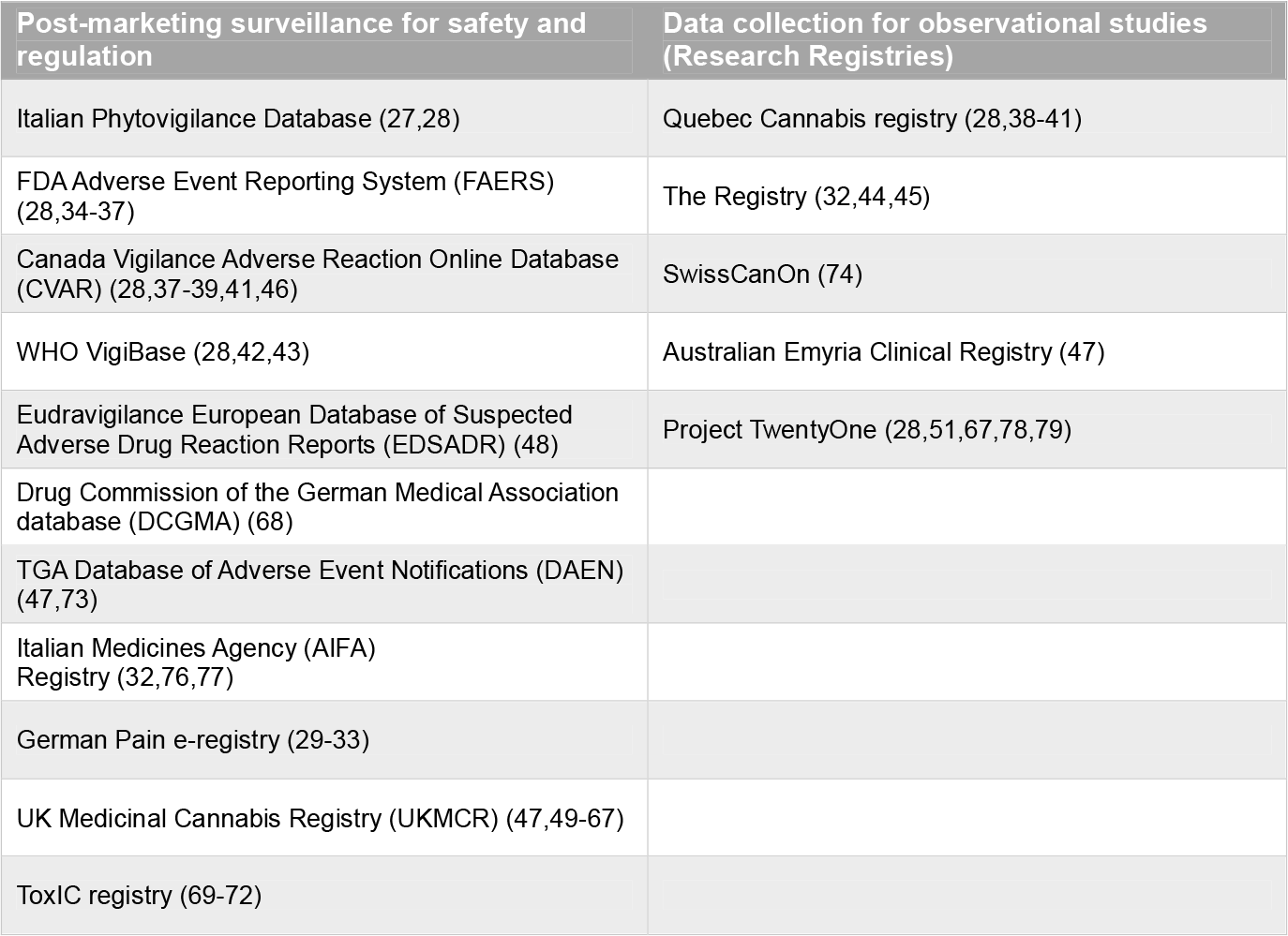
Primary purposes of monitoring systems for side effects and adverse events associated with CBM usage.

| Post-marketing surveillance for safety and regulation | Data collection for observational studies (Research Registries) |
| --- | --- |
| Italian Phytovigilance Database (27,28) | Quebec Cannabis registry (28,38-41) |
| FDA Adverse Event Reporting System (FAERS) (28,34-37) | The Registry (32,44,45) |
| Canada Vigilance Adverse Reaction Online Database (CVAR) (28,37-39,41,46) | SwissCanOn (74) |
| WHO VigiBase (28,42,43) | Australian Emyria Clinical Registry (47) |
| Eudravigilance European Database of Suspected Adverse Drug Reaction Reports (EDSADR) (48) | Project TwentyOne (28,51,67,78,79) |
| Drug Commission of the German Medical Association database (DCGMA) (68) |  |
| TGA Database of Adverse Event Notifications (DAEN) (47,73) |  |
| Italian Medicines Agency (AIFA) Registry (32,76,77) |  |
| German Pain e-registry (29-33) |  |
| UK Medicinal Cannabis Registry (UKMCR) (47,49-67) |  |
| Toxic registry (69-72) |  |

#### 2. Duration of data collection

All systems with post-marketing surveillance as a primary outcome, provide ongoing data collection at the time of our literature search.

Research registries, due to the longitudinal nature of observational studies, are also ongoing forms of monitoring, with two exceptions (Table 3). The Quebec Cannabis Registry, established in 2015, ceased data collection in 2018. Serious adverse events were reported to the Canada Vigilance Database for evaluation (37-40). The Registry collected data in the UK from 2012 until 2015, however data collection in Germany and Switzerland remains ongoing (31,43,44).

**Table 3.** Elements assessed by each monitoring system for side effects and adverse events associated with CBM usage.

|  | Formulation:<br>THC/CBD | Variety of<br>formulations | Dosage | Form/route of<br>administration | Patient demographics<br>mentioned | Indication | Effect | Concomitant<br>medication | Severity of<br>adverse reaction |
| --- | --- | --- | --- | --- | --- | --- | --- | --- | --- |
| Italian Phytovigilance Database (27,28)* | Yes | Yes | No | Yes | Age, weight, height, gender, comorbidities, pregnancy status | Yes | No | Yes | Yes |
| DAEN (47,73)* | Yes | Yes | Yes | Yes | Age, sex, gender, weight, ethnicity | Yes | No | Yes | Yes |
| WHO VigiBase (28,42,43)* | Yes | Yes | Yes | Yes | All relevant patient demographics as reported in case reports submitted via national regulatory authorities | Yes | No | Yes | Yes |
| Canada Vigilance Adverse Reaction Online Database (CVAR) (28,37-39,41,46)* | Yes | Yes | Yes | Yes | Age, sex, height, weight | Yes | No | Yes | Yes |
| Eudravigilance European Database of Suspected Adverse Drug Reaction Reports (EDSADR) (48)* | Yes | Yes | Yes | Yes | All relevant patient demographics as reported in case reports submitted via national regulatory authorities | Yes | No | Yes | Yes |
| Drug Commission of the German Medical Association database (DCGMA) (68)** | Not explicitly stated by study | Not explicitly stated by study | Not explicitly stated by study | Not explicitly stated by study | Not explicitly stated by study | Not explicitly stated by study | No | Not explicitly stated by study | Not explicitly stated by study |
| TGA Database of Adverse Event Notifications (DAEN) (47,73)** | Yes | Yes | Yes | Yes | Age, sex, height, weight, ethnicity, comorbidities maternal/paternal or foetal exposure | Yes | Yes | Yes | Yes |
| German Pain e-registry (29-33) | Not explicitly specified | Yes | Not explicitly stated | Not explicitly specified | Age, comorbidities | Yes | Yes | Yes | No |
| Quebec Cannabis registry (28,38-41) | Yes | Yes | Yes | Yes | Age, sex, occupation, comorbidities, smoking status, alcohol use and recreational drug use (at baseline), pregnancy/breastfeeding status, history of cannabis or substance use disorder | Yes | Yes | Yes | Yes |
| The Registry (32,44,45) | Yes (THC:CBD , nabiximols ) | No | Yes | Yes | Age, sex | Yes | Yes | Yes | Clinical significance determined by prescriber's professional opinion |
| Australian Emyria Clinical Registry (47) | Yes | Yes: all TGA approved CBMs | Yes | Oral | Age, comorbidities | Yes | Yes | Yes | Yes |
| UK Medicinal Cannabis Registry (UKMCR) (47,49-67) | Yes | Yes | Yes | Yes | Demographic, BMI, Comorbidities, drug, and alcohol history | Yes | Yes | Yes | Yes |
| ToxIC registry (69-72) | Yes | Yes, not cannabis-specific | Yes, a part of clinical data | Yes, part of clinical data | Demographics available in hospital patient records | Yes | No | Yes, a part of clinical data | Yes, mortality and whether life support was withdrawn signs and symptoms |
| SwissCanOn (74) | Not explicitly specified | Yes | Yes | Not explicitly specified | No mention of patient demographics | Yes, oncology | Yes | Not explicitly mentioned | Not explicitly mentioned |
| Italian Medicines Agency (AIFA) Registry (32,76,77) | Yes (THC: CBD, nabiximols, Sativex) | No, e-registry explicitly set up for Sativex reimbursement | Yes | Yes | Age, comorbidities | Yes, spasticity in multiple sclerosis | Yes | Yes, other use of antispastic drug | Yes |
| Project TwentyOne (28,51,67,78,79) | Yes | Yes | Not explicitly stated | Yes | Age, comorbidities | Yes: chronic pain, PTSD, anxiety, MS, Tourette's syndrome, Cannabis use disorder | Yes | Yes | Not explicitly stated |

\*Where primary reporting forms were accessible
\*\*Analysis for the purposes of this study were limited by availability of information

#### 3. Level of detail assessed

Most systems collect information on formulation and dosage of the CBM, however this is more common in regulatory databases. Route of administration and concomitant medications are frequently accounted for. Differences in details exist in assessing patient demographics. Age and sex of consumers are commonly collected; however, comorbidities and pregnancy status are not routinely reported (Table 3)

#### 4. Mode of monitoring: Spontaneous or mandatory?

Where data sources elect to participate in data collection, the system is considered to adopt a spontaneous reporting protocol. Therefore, all research registries, as well as the ToxiC registry, are spontaneous reporting systems (Table 3). The AIFA registry mandates reporting from patients. The German Pain E-registry, collects information from 200 pain centres across Germany, and fulfils the obligatory requirements of physicians to document patients under treatment for chronic pain (29-33).

#### 5. Reporter nature

Monitoring systems collect data from combinations of the following subgroups of individuals: patients, healthcare professionals, and/or manufacturers. Three registries (Table 4) utilise patient-reported outcomes. Four registries accept reports from healthcare professionals alone. The ToxiC registry (69-72), collecting data from medical records in participating hospitals, is included in this category. Larger databases encourage reports from patients, healthcare professionals, as well as manufacturers and producers of CBMs.

**Table 4.** Nature of reporters to each monitoring system for the side effects and adverse events associated with CBM usage.

| Patients only | Clinician/healthcare professionals only | Patients and healthcare professionals | Patients, healthcare professionals and manufacturers |
| --- | --- | --- | --- |
| Quebec Cannabis Registry (28,38-41) | The Registry (32,44,45) | Italian Phytovigilance Database (27,28) | FDA Adverse Event Reporting System (FAERS) (28,34-37) |
| SwissCanOn (74) | ToxiC Registry (69-72) | German Pain e-registry (29-33) | WHO VigiBase (28,42,43) |
| Project TwentyOne (29,52,68,78,79) | Drug Commission of the German Medical Association database (DCGMA) (68) | Australian Emyria Clinical Registry (47) | Canada Vigilance Adverse Reaction Online Database (28,37-39,41,46) |
| N/A | Italian Medicines Agency (AIFA) Database (32,76,77) | UK Medicinal Cannabis Registry (UKMCR) (47,49-67) | Eudravigilance European Database of Suspected Adverse Drug Reaction Reports (EDSADR) (48) |
| N/A | N/A | N/A | TGA Database of Adverse Event Notifications (DAEN) (47,73) |

#### 6. Specificity to cannabis vs other pharmaceuticals

Six of the sixteen monitoring systems captured by the search are specific to CBM monitoring. Databases offer assessments of adverse events associated with regulated and/or unregulated products within a region, rather than specific CBM monitoring.

#### 7. Affiliations

Four registries are affiliated with independent ownership (Table 5).

**Table 5.**
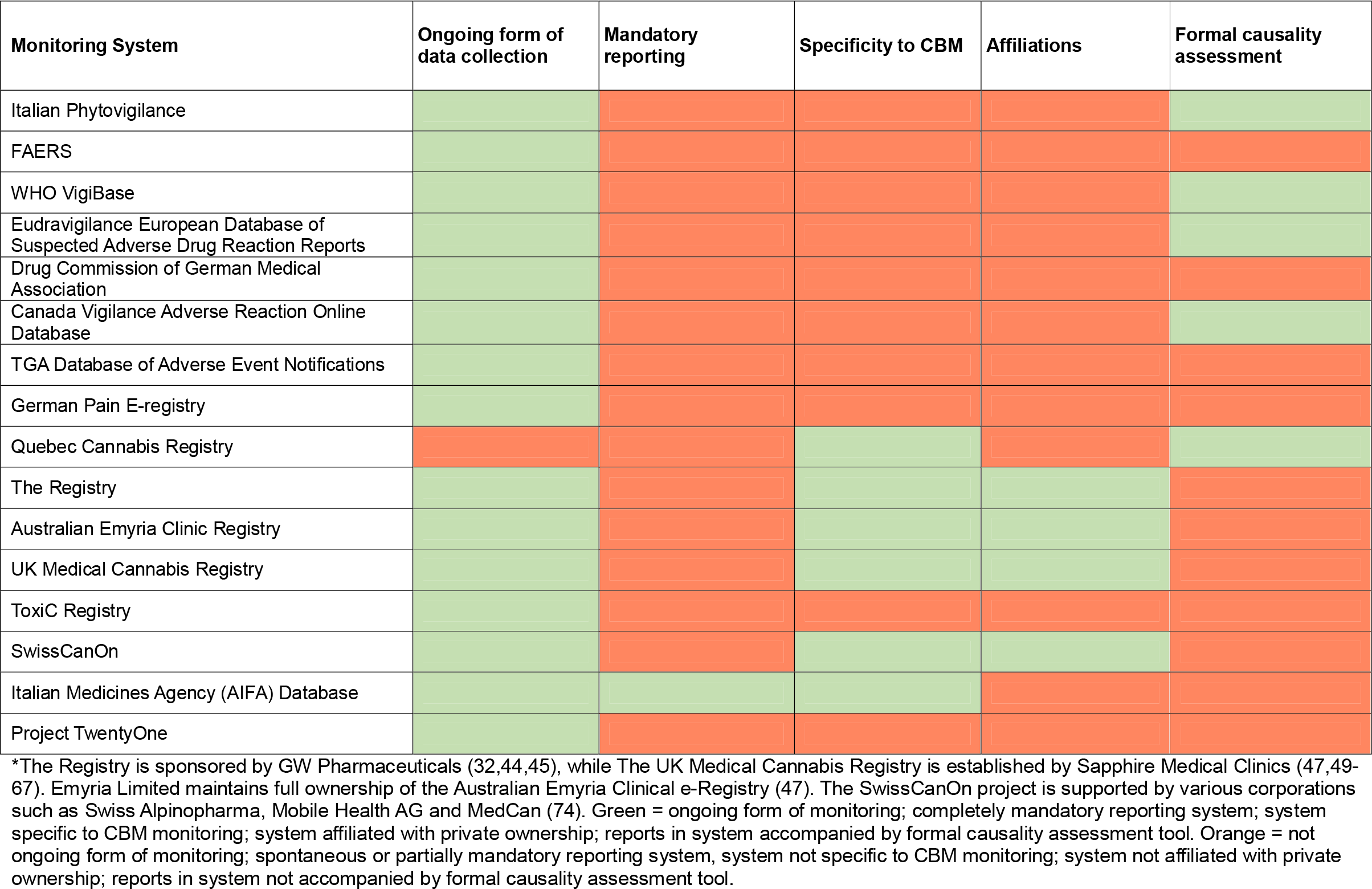
Features of monitoring systems for monitoring of side effect and adverse events associated with CBM usage*.

#### 8. Causality assessment

The strength of the causal relationship between CBM usage and the observed adverse event is considered in five monitoring systems (Table 5). The Quebec Cannabis Registry conducts causality assessments on reports, however the mode of assessment is not described in literature captured by our search (28,38-41). The Australian Emyria database does not implement a formal causality assessment, however possible causal relationships are guided by clinicians’ medical judgement (47).

## DISCUSSION

As of June 2023, there remains no robust and rigorous monitoring systems globally for collection of post-marketing safety data to accompany the international expansion in CBM uptake. The existence of several regulatory databases and multiple smaller registries globally, some of which have limited published data, demonstrates heterogeneity in post-marketing surveillance of CBM.

In some countries, the monitoring of CBM-related adverse events is embedded within the national regulatory framework for pharmaceuticals. The Italian Phytovigilance database, coordinated by the Italian National Institute of Health, collects reports on suspected adverse events associated with plant ingredient preparations and food supplements in Italy (81). Similarly, CVAR evaluates reports of suspected adverse reactions related to heath products with marketing authority within Canada (28). FAERS in the United States and DAEN in Australia follow similar frameworks (73,82). EDSADR, collecting data on suspected adverse reactions to authorized medicines or products undergoing trials in the European Economic Area, receives reports from National Medicine Regulatory Authorities and Marketing Authorization Holders that are submitted by patients and healthcare professionals (48). Many of these national databases input individual case reports into Vigibase (Supplementary Material 7), the database established by the WHO Programme for International Drug Monitoring, which bears greatest resemblance to a centralised monitoring system for CBM-related adverse events (28,42,43,83).

Other countries have established registries, either for observational studies or for regulatory purposes. Although some of these systems provide data targeted to cannabis, many rely on spontaneous reporting, and are therefore subject to selection bias from affiliations.

### Mandatory Vs Spontaneous Reporting Systems

The AIFA e-registry, to our knowledge, remains the only completely mandatory reporting system captured by our search, obligating patients to submit any side effects and adverse events experienced. The Italian Medicines Agency, under a reimbursement scheme called the Managed Entry Agreement, established an e-registry for all patients commencing on Sativex, to identify “non-responders” for subsequent reimbursement and discontinuation of treatment (76). EDSADR mandates Marketing Authorization Holders and National Competent Authorities to submit reports of adverse events received from patients and healthcare professionals. However, patients and healthcare professionals are not required to report adverse events (48). CVAR and TGA follow a similar reporting structure to EDSADR (28,73). Similarly, German pharmacists are obliged to report encountered suspected adverse reactions to the Drug Commission of the German Medical Association, however consumers are not required to report of side effects and adverse events (68).

The ad hoc nature of reporting requirements in many databases and registries risks a variety of reporting biases (84,85). These include underreporting, notoriety bias, and the preference to only report severe, usually rare, adverse events (84). Spontaneous adverse drug reaction reporting typically peaks following the second year of marketing, then subsequent declines, unaccompanied by changes in drug usage or adverse event incidence (86,87). Underreporting can be secondary to complacency, where adverse events are (incorrectly) believed to have been already well-documented following marketing. Uncertainty surrounding causal relationships is a further contributing factor to underreporting in spontaneous systems. Additionally, fear of medicolegal consequences, alongside overall clinician indifference is known to further discourage consistent reporting (85,88,89).

In November 2017, the change from spontaneous to mandatory submission of suspected ADRs from Marketing Authorizaation Holders and National Competent Authorities to the Eudravigilance database, resulted in a significant increase in the number of reports collected (90). Therefore, mandatory reporting framework appear to mitigate issues of underreporting. Additional interventions such as financial incentives, training on ADR selection and existing reporting systems, as well as continuous feedback on safety signals identified may lessen other reporting biases intrinsic to spontaneous reporting systems.

### Causality assessment

Although post-marketing surveillance is more likely to identify a strong causal relationship between CBMs and adverse events, rather than certain proof of causality, there is variability in the level of causality assessment accepted by each monitoring system (91). The Italian Phytovigilance database and CVAR both use the WHO-UMC causality classification system, whereas many other registries and databases such as the FAERS and DAEN do not implement a formal causality assessment process. Within these systems, reports are broadly classified as monitoring “suspected” adverse events. Factors known to help determine the strength of a causal relationship include the temporal relationship between the commencement of CBM and adverse event onset, as well as response to ceasing the CBM and subsequent re-administration (87). The FAERS reporting form provides an opportunity for details on response to de-challenge and rechallenge with the drug agent, however these fields are not mandatory for report submission. VigiBase, collecting case reports from national pharmacovigilance centres internationally, notes discrepancies between reports and do not validate causality claims (27, 41-43). Eudravigilance similarly accepts reports of varying strengths of causality (48). A standardised method of determining causality across monitoring systems may improve the strength of safety data derived from reports of adverse events.

### Diversity in report quality and detail

There are further inconsistencies in the level of detail collected by each monitoring system. Some databases do not specify dosage or route of administration of the CBM, decreasing the utility of available data in determining accurate safety data. Information collected on patient demographics also varies significantly between systems, especially when considering a patient’s comorbidities, pregnancy status and concomitant medications. As potential confounding factors in determining the cause of the adverse reaction, variations in these demographic details impacts the interpreting of the safety data collected (92,93).

Variability in quality of reports poses an additional problem in centralised databases such as VigiBase, where data is derived from various sources. Between healthcare professionals, consumers, and manufacturers from over 150 countries (27,41,42,94), differences exist in terminology, coding practices and reporting standards across these different regions and healthcare systems. Standard terminology for drug reactions and medicinal products are codified using the Medical Dictionary for Regulatory Activities (MedDRA) and WHODrug classifications respectively (95). However, data at a national level may be mistranslated and inexact when transferred to the standardised VigiBase terminology. For instance, CBD-dominant cannabis products may be coded as CBD according to the standardised WHODrug system, when their THC content would qualify them to be coded as *Cannabis Sativa* whole extracts. The current MedDRA classification for severity of adverse reactions also provides limited descriptors for cannabis-related adverse events, such as ‘cannabis hyperemesis syndrome’ or ‘cannabis dependence’ and ‘withdrawal’ (28). Such discrepancies limit data comparability and complicate assessment of adverse events associated with CBM usage.

The diversity of data sources reporting to a centralised system such as VigiBase predisposes to potential duplication reports. Although efforts are made to identify duplicates, slight differences in nomenclature of related reports may allow duplicates to bypass the algorithm employed by VigiBase (95). As such, the development of one central reporting system with standardised nomenclature and formatting may help with the imprecision in adapting multiple data sources into a standardised framework.

### Affiliations

The affiliation of certain registries with independent ownership presents risk of selection bias when evaluating results. The Registry, a multicentre observational research registry collecting data from UK, Germany, and Switzerland, is sponsored by GW Pharmaceuticals, the manufacturers of the THC: CBD oromucosal spray (Nabiximol). Prescribers were identified and invited to participate in data collection by GW Pharmaceuticals, and nominally compensated for completing Case Report Forms (44). UKMCR is maintained by Sapphire Medical Clinics, inviting patients from a private healthcare setting not representative of the broader population of CBM consumers (47,49-67). Similarly, the Australian Emyria Clinical e-Registry sources participation from Emerald Clinics, a network of clinics specialising in use of currently unregistered medicines and commercialisation of collected clinical evidence with Spectrum Therapeutics, the medical division of a cannabis company known as Canopy Growth (47,96). Of note, patients from these registries are a specific subset of CBM consumers, and these registries are aligned with various companies that have interests outside that of the accumulation of real-world safety data.

### Accessibility to reporting and safety data

Accessibility to reporting forms, time constraints, and awareness of existing reporting schemes have been forwarded as factors limiting participation in monitoring systems (97-99). Additionally, there exists a delay between onset and recognition of the adverse drug reaction, and another lag between reports being input into national pharmacovigilance centres and successful transfer into a central monitoring system such as VigiBase (95). Between access issues to reporting forms, as well as access to safety data published by databases, current monitoring systems are difficult to incorporate into the busy workflow of clinical practice. As such, monitoring of CBM adverse events, from reporting to publishing of safety data, requires a streamlined approach to parallel speed of CBM uptake.

## LIMITATIONS

Information on details collected in smaller registries is limited by the availability of published data, as often the original reporting form was inaccessible via a secondary Google Search. Additionally, the discipline of pharmacovigilance posits a difference between the definition of side effects, adverse events, and adverse reactions (100). However, these terms were used interchangeably in papers, and therefore adverse events were assumed to encompass side effects. Given the international scope of our data collection, our paper was further limited by the exclusion of papers not published in the English language.

## CONCLUSION

To the best of our knowledge, this is the first scoping review assessing the existing monitoring systems for side effects and adverse events associated with medicinal cannabis usage at an international level. As a novel therapeutic, CBM may be a promising solution for an increasing range of intractable conditions. Our narrative review with a systematic approach has identified various issues with the quality, access, consistency, and attitudes towards existing reporting systems for monitoring of adverse events related to CBM usage. Although the ideal international monitoring system has proven difficult amongst the evolving landscape of cannabis legalisation, there still remains a key need for a centralised, and standardised system, that is accessible and operates in real time. Post-marketing safety data captured in this way, accompanying the growth in clinical use, will support both public and clinical interest in CBMs as a therapeutic in a safe and efficient manner.

## Data Availability

All data produced in the present work are contained in the manuscript

## LIST OF ABBREVIATIONS

CBM: Cannabis-based medicines
TGA: Therapeutic Goods’ Administration
CBD: Cannabidiol
THC: Delta-9-tetrahydrocannabinol
MS: Multiple sclerosis
RA: Rheumatoid arthritis Irritable Bowel syndrome: IBS
FAERS: FDA Adverse events reporting system
CVAR: Canada Vigilance Adverse Reaction Online Database
EDSADR: Eudravigilance European Database of Suspected Adverse Drug Reaction Reports
DAEN: TGA Database of Adverse event Notifications
UKMCR: UK Medical Cannabis Registry
AIFA: Italian Medicines Agency

## ACKNOWLEDGEMENTS

Thank you to my Doctor of Medicine supervisors, Dr Christine Hallinan and A/Prof Yvonne Bonomo from the University of Melbourne for their support and encouragement in completing this review.

## REFERENCES

1. Russo EB. History of cannabis and its preparations in saga, science, and sobriquet. Chemistry & biodiversity. 2007;4(8):1614–1648.

2. Bridgeman MB, Abazia DT. Medicinal Cannabis: History, Pharmacology, And Implications for the Acute Care Setting. P t. Mar 2017;42(3):180–188.

3. Shover CL, Humphreys K. Six policy lessons relevant to cannabis legalization. Am J Drug Alcohol Abuse. 2019;45(6):698–706. 10.1080/00952990.2019.1569669

4. Baratta F, Simiele M, Pignata I, et al. Cannabis-Based Oral Formulations for Medical Purposes: Preparation, Quality and Stability. Pharmaceuticals (Basel). Feb 22 2021;14(2)10.3390/ph14020171

5. Hazekamp A, Ware MA, Muller-Vahl KR, Abrams D, Grotenhermen F. The Medicinal Use of Cannabis and Cannabinoids—An International Cross-Sectional Survey on Administration Forms. Journal of Psychoactive Drugs. 2013/07/01 2013;45(3):199–210. 10.1080/02791072.2013.805976

6. National Academies of Sciences E, Medicine, Health, et al. The National Academies Collection: Reports funded by National Institutes of Health. The Health Effects of Cannabis and Cannabinoids: The Current State of Evidence and Recommendations for Research. National Academies Press (US) Copyright 2017 by the National Academy of Sciences. All rights reserved.; 2017.

7. Arnold JC, Nation T, McGregor IS. Prescribing medicinal cannabis. Aust Prescr. Oct 2020;43(5):152–159. 10.18773/austprescr.2020.052

8. Hallinan CM, Gunn JM, Bonomo YA. Use of electronic medical records to monitor the safe and effective prescribing of medicinal cannabis: is it feasible? Article. Australian Journal of Primary Health. 2022;10.1071/PY22054

9. TGA. Medicinal Cannabis Access Data Dashboard. Australian Government Department of Health Accessed 07/12/2023, https://www.tga.gov.au/medicinal-cannabis-access-data-dashboard

10. TGA. Medicinal Cannabis Authorised Prescriber Scheme data. Australian Government Therapeutic Goods Administration (TGA) 2023. Accessed September 07, 2023. https://www.tga.gov.au/products/unapproved-therapeutic-goods/medicinal-cannabis-hub/medicinal-cannabis-access-pathways-and-patient-access-data/medicinal-cannabis-authorised-prescriber-scheme-data

11. Clarke H. The evolving culture of medical cannabis in Canada for the management of chronic pain. Frontiers in Pharmacology. April 7 2023;1410.3389/fphar.2023.1153584

12. Sznitman SR, Bretteville-Jensen AL. Public opinion and medical cannabis policies: examining the role of underlying beliefs and national medical cannabis policies. Harm reduction journal. 2015;12(1):1–10.

13. Wong SS, Wilens TE. Medical uses of cannabinoids in children and adolescents: A systematic review. Journal of the American Academy of Child and Adolescent Psychiatry. 2017;56(10):S295. 64th Annual Meeting American Academy of Child and Adolescent Psychiatry, AACAP 2017. Washington, DC United States. 10.1016/j.jaac.2017.09.399

14. National Academies of Sciences E, Medicine. The health effects of cannabis and cannabinoids: the current state of evidence and recommendations for research. 2017,

15. Bonomo Y, Souza JDS, Jackson A, Crippa JAS, Solowij N. Clinical issues in cannabis use. Br J Clin Pharmacol. Nov 2018;84(11):2495–2498. 10.1111/bcp.13703

16. Bonn-Miller MO, Loflin MJE, Thomas BF, Marcu JP, Hyke T, Vandrey R. Labeling Accuracy of Cannabidiol Extracts Sold Online. JAMA. 2017;318(17):1708–1709. 10.1001/jama.2017.11909

17. Hallinan C, Eden E, Graham M, … Over the counter low-dose cannabidiol: A viewpoint from the ACRE Capacity Building Group. Journal of …. 2022;10.1177/02698811211035394

18. Fitzcharles M-A, Shir Y, Häuser W. Medical cannabis: strengthening evidence in the face of hype and public pressure. Canadian Medical Association Journal. 2019;191(33):E907–E908. 10.1503/cmaj.190509

19. Page MJ, McKenzie JE, Bossuyt PM, et al. The PRISMA 2020 statement: An updated guideline for reporting systematic reviews. PLOS Medicine. 2021;18(3):e1003583. 10.1371/journal.pmed.1003583

20. Munn Z, Peters MDJ, Stern C, Tufanaru C, McArthur A, Aromataris E. Systematic review or scoping review? Guidance for authors when choosing between a systematic or scoping review approach. BMC Medical Research Methodology. 2018/11/19 2018;18(1):143. 10.1186/s12874-018-0611-x

21. Haddaway NR, Collins AM, Coughlin D, Kirk S. The Role of Google Scholar in Evidence Reviews and Its Applicability to Grey Literature Searching. PLOS ONE. 2015;10(9):e0138237. 10.1371/journal.pone.0138237

22. Paez A. Gray literature: An important resource in systematic reviews. Journal of Evidencel Based Medicine. 2017;10(3):233–240.

23. Adams J, Hillier-Brown FC, Moore HJ, et al. Searching and synthesising ‘grey literature’ and ‘grey information’ in public health: critical reflections on three case studies. Systematic Reviews. 2016/09/29 2016;5(1):164. 10.1186/s13643-016-0337-y

24. Paut Kusturica M, Tomas A, Sabo A, Tomić Z, Horvat O. Medical cannabis: Knowledge and attitudes of prospective doctors in Serbia. Saudi Pharm J. Mar 2019;27(3):320–325. 10.1016/j.jsps.2018.11.014

25. Hallinan C, Gunn J, Bonomo Y. Implementation of medicinal cannabis in Australia: innovation or upheaval? Perspectives from physicians as key informants, a qualitative analysis. BMJ open. bmjopen.bmj.com; 2021.

26. Lipnik-Štangelj M, Razinger B. A regulatory take on cannabis and cannabinoids for medicinal use in the European Union. Arh Hig Rada Toksikol. Mar 1 2020;71(1):12–18. 10.2478/aiht-2020-71-3302

27. Crescioli G, Bettiol A, MennitiJIppolito F, et al. Adverse events following cannabis for medical use in Tuscany: An analysis of the Italian Phytovigilance database. British Journal of Clinical Pharmacology. 2020;86(1):106–120. 10.1111/bcp.14140

28. Jack S. Pharmacovigilance of Cannabis Products for Medical and Non-medical Purposes. In: Barnes J, ed. Pharmacovigilance for Herbal and Traditional Medicines: Advances, Challenges and International Perspectives. Springer International Publishing; 2022. 10.1007/978-3-031-07275-8_20

29. Ueberall MA, Horlemann J, Schuermann N, Kalaba M, Ware MA. Effectiveness and Tolerability of Dronabinol Use in Patients with Chronic Pain: A Retrospective Analysis of 12-Week Open-Label Real-World Data Provided by the German Pain e-Registry. Pain Med. Aug 1 2022;23(8):1409–1422. 10.1093/pm/pnac010

30. Ueberall MA, Essner U, Mueller-Schwefe GH. Effectiveness and tolerability of THC:CBD oromucosal spray as add-on measure in patients with severe chronic pain: analysis of 12-week open-label real-world data provided by the German Pain e-Registry. J Pain Res. 2019;12:1577–1604. 10.2147/jpr.S192174

31. Ueberall MA, Essner U, Silván CV, Mueller-Schwefe GH. Comparison of the Effectiveness and Tolerability of Nabiximols (THC:CBD) Oromucosal Spray versus Oral Dronabinol (THC) as Add-on Treatment for Severe Neuropathic Pain in Real-World Clinical Practice: Retrospective Analysis of the German Pain e-Registry. Journal of Pain Research. 2022;15:267–286. 10.2147/JPR.S340968

32. Prieto González JM, Vila Silván C. Safety and tolerability of nabiximols oromucosal spray: a review of real-world experience in observational studies, registries, and case reports. Expert Rev Neurother. May 2021;21(5):547–558. 10.1080/14737175.2021.1904896

33. Ueberall MA, Silván CV, Essner U, Mueller-Schwefe GH. Effectiveness, Safety, and Tolerability of Nabiximols Oromucosal Spray vs Typical Oral Long-Acting Opioid Analgesics in Patients with Severe Neuropathic Back Pain: Analysis of 6-Month Real-World Data from the German Pain e-Registry. Pain Medicine. Apr 2022 2022;23(4):745–760. 10.1093/pm/pnab263

34. Lunghi C, Fusaroli M, Giunchi V, Raschi E, Zongo A, Poluzzi E. What Is the Safety Profile of Cannabis-Based Medications? Analysis of the Post-Marketing Signals from the FDA Adverse Event Reporting System. Drug Safety. 2022;45(10):1242–1243. 21st Annual Meeting of the International Society of Pharmacovigilance, ISoP 2022. Verona Italy. 10.1007/s40264-022-01219-7

35. Simon TA, Simon JH, Heaning EG, Gomez-Caminero A, Marcu JP. Delta-8, a Cannabis-Derived Tetrahydrocannabinol Isomer: Evaluating Case Report Data in the Food and Drug Administration Adverse Event Reporting System (FAERS) Database. Drug, Healthcare and Patient Safety. 2023;15:25–38. 10.2147/DHPS.S391857

36. Piracha Z, Ramnarain R. Analyzing Medwatch Data in an Effort to Assess Dronabinol Post-Marketing Safety. Current Trends in Biomedical Engineering & Biosciences. 2017;3(4):64–65. https://ideas.repec.org/a/adp/jctbeb/v3y2017i4p64-65.html

37. Sharma V, Gelin LFF, Sarkar IN. Identifying Herbal Adverse Events From Spontaneous Reporting Systems Using Taxonomic Name Resolution Approach. Article. Bioinformatics and Biology Insights. 2020;1410.1177/1177932220921350

38. Aprikian S, Kasvis P, Vigano M, Hachem Y, Canac-Marquis M, Vigano A. Medical cannabis is effective for cancer-related pain: Quebec Cannabis Registry results. BMJ Supportive and Palliative Care. 2023;10.1136/spcare-2022-004003

39. Moride Y, Hachem Y, Castilloux AM, et al. Oral Presentation: Safety of Medical Cannabis: A Descriptive Study Using the Quebec Cannabis Registry [Abstract]. Drug Safety. Springer International Publishing; 2022:1128–1129. 21st Annual Meeting of the International Society of Pharmacovigilance, ISoP 2022. Verona Italy.; vol. 10.

40. Vigano A, Canac-Marquis M, Gamaoun R, et al. The Quebec Cannabis Registry: a pharmacovigilance and effectiveness study on the use of medical cannabis in cancer patients. Journal of Clinical Oncology. 2020;38(15):12109–12109. 10.1200/JCO.2020.38.15_suppl.12109

41. Vigano A, Moride Y, Hachem Y, et al. The Quebec Cannabis Registry: Investigating the Safety and Effectiveness of Medical Cannabis. Cannabis and cannabinoid research. 2022;10.1089/can.2022.0041

42. van--Hunsel F, Gattepaille LM, Westerberg C, Barnes J. Reports for herbal medicines in the global suspected ADR database VigiBase. Pharmacovigilance for Herbal and Traditional Medicines: Advances, Challenges and International Perspectives. Springer; 2022.

43. Pochet S, Lechon AS, Lescrainier C, et al. Herb-anticancer drug interactions in real life based on VigiBase, the WHO global database. Scientific Reports. 2022;12(1)14178. 10.1038/s41598-022-17704-z

44. Etges T, Karolia K, Grint T, et al. An observational postmarketing safety registry of patients in the UK, Germany, and Switzerland who have been prescribed Sativex(®) (THC:CBD, nabiximols) oromucosal spray. Ther Clin Risk Manag. 2016;12:1667–1675. 10.2147/tcrm.S115014

45. Oreja-Guevara C, Casanova B, Ordás C, Asensio D. Observational safety study of THC: CBD oromucosal spray (Sativex) in multiple sclerosis patients with spasticity. Clin Exp Pharmacol. 2015;5(184)10.4172/2161-1459.1000184

46. Jack S, Huff SP, Abramovici H. Inter-Year Comparison of Adverse Reactions Associated with Legal Cannabis Products Reported to Health Canada, 2018-2019 to 2021. Drug Safety. Springer International Publishing; 2022:1313. 21st Annual Meeting of the International Society of Pharmacovigilance, ISoP 2022. Verona Italy.; vol. 10.

47. Vickery AW, Roth S, Ernenwein T, Kennedy J, Washer P. A large Australian longitudinal cohort registry demonstrates sustained safety and efficacy of oral medicinal cannabis for at least two years. PLoS One. 2022;17(11)10.1371/journal.pone.0272241

48. Ammendolia I, Mannucci C, Cardia L, Gangemi S, Esposito E, Calapai F. Pharmacovigilance on cannabidiol as an antiepileptic agent. Frontiers in Pharmacology. 2023;14.10.3389/fphar.2023.1091978

49. Erridge S, Salazar O, Kawka M, et al. An initial analysis of the UK Medical Cannabis Registry: Outcomes analysis of first 129 patients. Neuropsychopharmacol Rep. 2021;41(3):362–370. 10.1002/npr2.12183

50. Olsson F, Erridge S, Tait J, et al. An observational study of safety and clinical outcome measures across patient groups in the United Kingdom Medical Cannabis Registry. Expert Review of Clinical Pharmacology. 2023;16(3):257–266. 10.1080/17512433.2023.2183841

51. Ergisi M, Erridge S, Harris M, et al. An Updated Analysis of Clinical Outcome Measures Across Patients from the UK Medical Cannabis Registry. Cannabis and Cannabinoid Research. 2023;8(3):557–566. 10.1089/can.2021.0145

52. Wang C, Erridge S, Holvey C, et al. Assessment of clinical outcomes in patients with fibromyalgia: Analysis from the UK Medical Cannabis Registry. Brain and Behavior. 2023;13(7)10.1002/brb3.3072

53. Pillai M, Erridge S, Bapir L, et al. Assessment of clinical outcomes in patients with post-traumatic stress disorder: analysis from the UK Medical Cannabis Registry. Expert Review of Neurotherapeutics. 2022;22(11):1009–1018. 10.1080/14737175.2022.2155139

54. Mangoo S, Erridge S, Holvey C, et al. Assessment of clinical outcomes of medicinal cannabis therapy for depression: analysis from the UK Medical Cannabis Registry. Expert Review of Neurotherapeutics. 2022;22(11):995–1008. 10.1080/14737175.2022.2161894

55. Erridge S, Kerr-Gaffney J, Holvey C, et al. Clinical outcome analysis of patients with autism spectrum disorder: analysis from the UK Medical Cannabis Registry. Therapeutic Advances in Psychopharmacology. 2022;1210.1177/20451253221116240

56. Rifkin-Zybutz R, Erridge S, Holvey C, et al. Clinical outcome data of anxiety patients treated with cannabis-based medicinal products in the United Kingdom: a cohort study from the UK Medical Cannabis Registry. Psychopharmacology. 2023;240(8):1735–1745. 10.1007/s00213-023-06399-3

57. Erridge S, Holvey C, Coomber R, et al. Clinical Outcome Data of Children Treated with Cannabis-Based Medicinal Products for Treatment Resistant Epilepsy-Analysis from the UK Medical Cannabis Registry. Neuropediatrics. 2022;54(3):174–181. 10.1055/a-2002-2119

58. Tait J, Erridge S, Holvey C, et al. Clinical outcome data of chronic pain patients treated with cannabis-based oils and dried flower from the UK Medical Cannabis Registry. Expert Review of Neurotherapeutics. 2023;23(4):413–423. 10.1080/14737175.2023.2195551

59. Kawka M, Erridge S, Holvey C, et al. Clinical Outcome Data of First Cohort of Chronic Pain Patients Treated With Cannabis-Based Sublingual Oils in the United Kingdom: Analysis From the UK Medical Cannabis Registry. Journal of Clinical Pharmacology. 2021;61(12):1545–1554. 10.1002/jcph.1961

60. Dalavaye N, Erridge S, Nicholas M, et al. The effect of medical cannabis in inflammatory bowel disease: analysis from the UK Medical Cannabis Registry. Expert Review of Gastroenterology and Hepatology. 2023;17(1):85–98. 10.1080/17474124.2022.2161046

61. Tait J, Erridge S, Sodergren MH. UK Medical Cannabis Registry: A Patient Evaluation. Journal of Pain and Palliative Care Pharmacotherapy. 2023;37(2):170–177. 10.1080/15360288.2023.2174633

62. Ergisi M, Erridge S, Harris M, et al. UK Medical Cannabis Registry: an analysis of clinical outcomes of medicinal cannabis therapy for generalized anxiety disorder. Expert Review of Clinical Pharmacology. 2022;15(4):487–495. 10.1080/17512433.2022.2020640

63. Nicholas M, Erridge S, Bapir L, et al. UK medical cannabis registry: assessment of clinical outcomes in patients with headache disorders. Expert Review of Neurotherapeutics. 2023;23(1):85–96. 10.1080/14737175.2023.2174017

64. Holvey C, Coomber R, Hoare J, et al. Clinical Outcome Data of Children Treated with Cannabis-Based Medicinal Products for Treatment Resistant Epilepsy—Analysis from the UK Medical Cannabis Registry. Neuropediatrics. January 30 2023;54(3):174–181. 10.1055/a-2002-2119

65. Harris M, Erridge S, Ergisi M, et al. UK Medical Cannabis registry: an analysis of clinical outcomes of medicinal cannabis therapy for chronic pain conditions. Expert Review of Clinical Pharmacology. 2022;14(4):473–485. 10.1080/17512433.2022.2017771

66. Nimalan D, Kawka M, Erridge S, et al. UK Medical Cannabis Registry palliative care patients cohort: initial experience and outcomes. Journal of Cannabis Research. 2022;4(1):33. 10.1186/s42238-021-00114-9

67. Lynskey M, Fayaz A, Athanasiou-Fragkouli A, et al. Characteristics of People Seeking Prescribed Cannabinoids for the Treatment of Chronic Pain: Evidence From Project Twenty 21. Frontiers in Pain Research. June 14 2022;310.3389/fpain.2022.891498

68. Freudewald L, Iliescu O, Robert NP, AndreSaid A, Schulz M. Medicinal Cannabis and Related Products-Analyses of Quality Defects and Adverse Drug Reactions Reported by German Community Pharmacists [Abstract]. Drug Safety. Springer International Publishing; 2022:1240–1241. 21st Annual Meeting of the International Society of Pharmacovigilance, ISoP 2022. Verona Italy.; vol. 10.

69. Spyres MB, Ruha AM, Wax PM, Brent J. Clinical and demographic factors in marijuana toxicity: The ToxIC registry experience since 2010. Clinical Toxicology. 2015;53(7):677. 10.3109/15563650.2015.1071025

70. Spyres MB, Farrugia LA, Kang AM, et al. The Toxicology Investigators Consortium Case Registry-the 2019 Annual Report. Journal of Medical Toxicology. 2020;16(4):361–387. 10.1007/s13181-020-00810-7

71. Love JS, Karshenas DL, Spyres MB, et al. The Toxicology Investigators Consortium Case Registry-the 2021 Annual Report. Journal of Medical Toxicology. 2022;18(4):267–296. 10.1007/s13181-022-00910-6

72. Rhyee SH, Campleman SL, Judge B, et al. The Toxicology Investigators Consortium Case Registry—the 2017 Annual Report. Journal of Medical Toxicology. 2018;14(3):182–211. 10.1007/s13181-018-0679-z

73. TGA. Database of Adverse Event Notifications (DAEN) Updated July 12, 2023. Accessed August 13, 2023. https://www.tga.gov.au/safety/safety/safety-monitoring-daen-database-adverse-event-notifications/database-adverse-event-notifications-daen

74. Trojan A, Breitkopf S, Pittl S, Heeren M. SwissCanOn - scientific patient registry for medicinal cannabis in oncology including ePROs - Trial in Progress [Abstract]. 5th Swiss Oncology and Hematology Congress; Basel Switzerland. EMH Swiss Medical Publishers Ltd.; 2022:chap 265. https://smw.ch/fileadmin/content/supplements/SMW-152-40034.pdf http://ovidsp.ovid.com/ovidweb.cgi?T=JS&PAGE=reference&D=emexa&NEWS=N&AN=639854642

75. Mahabir VK, Merchant JJ, Smith C, Garibaldi A. Medical cannabis use in the United States: a retrospective database study. J Cannabis Res. Sep 29 2020;2(1):32. 10.1186/s42238-020-00038-w

76. Messina S, Solaro C, Righini I, et al. Sativex in resistant multiple sclerosis spasticity: Discontinuation study in a large population of Italian patients (SA.FE. study). PLoS One. 2017;12(8)10.1371/journal.pone.0180651

77. Patti F, Messina S, Solaro C, et al. Efficacy and safety of cannabinoid oromucosal spray for multiple sclerosis spasticity. Journal of Neurology, Neurosurgery and Psychiatry. 2016;87(9):944–951. 10.1136/jnnp-2015-312591

78. O’Brien K, Beilby J, Frans M, et al. Preliminary findings from project Twenty21 Australia: An observational study of patients prescribed medicinal Cannabis for chronic pain, anxiety, posttraumatic stress disorder and multiple sclerosis. Drug Science, Policy and Law. January 1 2023;910.1177/20503245231164718

79. Sakal C, Lynskey M, Schlag AK, Nutt DJ. Developing a real-world evidence base for prescribed cannabis in the United Kingdom: preliminary findings from Project Twenty21. Psychopharmacology (Berl). May 2022;239(5):1147–1155. 10.1007/s00213-021-05855-2

80. Aviram J, Pud D, Schiff-Keren B, et al. The Israeli Multi-Center Registry of Medical Cannabis (MC) for Chronic Pain: Current Findings. Rambam Maimonides Medical Journal. 2018;910.5041/RMMJ.10329

81. Menniti-Ippolito F, Firenzuoli F. The Italian Phytovigilance Spontaneous Reporting Scheme. In: Barnes J, ed. Pharmacovigilance for Herbal and Traditional Medicines: Advances, Challenges and International Perspectives. Springer International Publishing; 2022. 10.1007/978-3-031-07275-8_17

82. Pozsgai K, Szűcs G, Kőnig-Péter A, et al. Analysis of pharmacovigilance databases for spontaneous reports of adverse drug reactions related to substandard and falsified medical products: A descriptive study. Front Pharmacol. 2022;13:964399. 10.3389/fphar.2022.964399

83. WHO. VigiBase: Who’s global database signalling harm and pointing to safer use [Internet]. https://who-umc.org/vigibase/vigibase-who-s-global-database/

84. Matsuda S, Aoki K, Kawamata T, et al. Bias in Spontaneous Reporting of Adverse Drug Reactions in Japan. PLOS ONE. 2015;10(5):e0126413. 10.1371/journal.pone.0126413

85. Palleria C, Leporini C, Chimirri S, et al. Limitations and obstacles of the spontaneous adverse drugs reactions reporting: Two “challenging” case reports. J Pharmacol Pharmacother. Dec 2013;4(Suppl 1):S66–72. 10.4103/0976-500x.120955

86. Weber J. Epidemiology of adverse reactions to nonsteroidal antiJinflammatory drugs. Adv Inflammation Res. 1984;6.,

87. Goldman SA. Limitations and strengths of spontaneous reports data. Clinical Therapeutics. 1998;20.10.1016/S0149-2918(98)80007-6

88. Biagi C, Montanaro N, Buccellato E, Roberto G, Vaccheri A, Motola D. Underreporting in pharmacovigilance: an intervention for Italian GPs (Emilia–Romagna region). European journal of clinical pharmacology. 2013;69:237–244.

89. Costa C, Abeijon P, Rodrigues DA, Figueiras A, Herdeiro MT, Torre C. Factors associated with underreporting of adverse drug reactions by patients: a systematic review. International Journal of Clinical Pharmacy. May 29 2023;10.1007/s11096-023-01592-y

90. Candore G, Monzon S, Slattery J, et al. The Impact of Mandatory Reporting of Non-Serious Safety Reports to EudraVigilance on the Detection of Adverse Reactions. Drug Saf. Jan 2022;45(1):83–95. 10.1007/s40264-021-01137-0

91. Hammad TA, Afsar S, McAvoy LB, Le Louet H. Aspects to consider in causality assessment of safety signals: broadening the thought process. Methods. Frontiers in Drug Safety and Regulation. 2023-May-25 2023;310.3389/fdsfr.2023.1193413

92. Gottschling S, Ayonrinde O, Bhaskar A, et al. Safety considerations in cannabinoid-based medicine. International Journal of General Medicine. 2020;13:1317–1333. 10.2147/IJGM.S275049

93. Grzeskowiak LE, Grieger JA, Andraweera P, et al. The deleterious effects of cannabis during pregnancy on neonatal outcomes. Medical Journal of Australia. 2020;212(11):519–524.

94. Lindquist M. VigiBase, the WHO Global ICSR Database System: Basic Facts. Drug Information Journal. 2008/09/01 2008;42(5):409–419. 10.1177/009286150804200501

95. WHO. Guideline for using Vigibase data in studies. Uppsula Monitoring Centre. Accessed September, 2023. https://who-umc.org/media/05kldqpj/guidelineusingvigibaseinstudies.pdf

96. Tchetvertakov G. Emerald Clinics signs cannabis RWE contract with Canopy Growth’s UK subsidiary Spectrum Biomedical. Accessed September, 2023. https://smallcaps.com.au/emerald-clinics-cannabis-rwe-canopy-growth-uk-subsidiary-spectrum-biomedical/

97. Herdeiro MT, Figueiras A, Polónia J, Gestal-Otero JJ. Physicians’ Attitudes and Adverse Drug Reaction Reporting. Drug Safety. September 1 2005;28(9):825–833. 10.2165/00002018-200528090-00007

98. Belton KJ, Lewis SC, Payne S, Rawlins M, Wood S. Attitudinal survey of adverse drug reaction reporting by medical practitioners in the United Kingdom British journal of clinical pharmacology. 1995;39(3):223–226.

99. Bäckström M, Mjörndal T, Dahlqvist R, Nordkvist-Olsson T. Attitudes to reporting adverse drug reactions in northern Sweden. European journal of clinical pharmacology. 2000;56:729–732.

100. World Health Organization. Quality A, Safety of Medicines T. Safety of medicines : a guide to detecting and reporting adverse drug reactions : why health professionals need to take action. Geneva: World Health Organization; 2002.

