## Supplementary Material for "An overview of global monitoring systems for the side effects and adverse events associated with medicinal cannabis use: A scoping review using a systematic approach"

**Supplementary 1. Number of Patients newly prescribed Medicinal Cannabis as reported by TGA's Authorised Prescriber Scheme.**

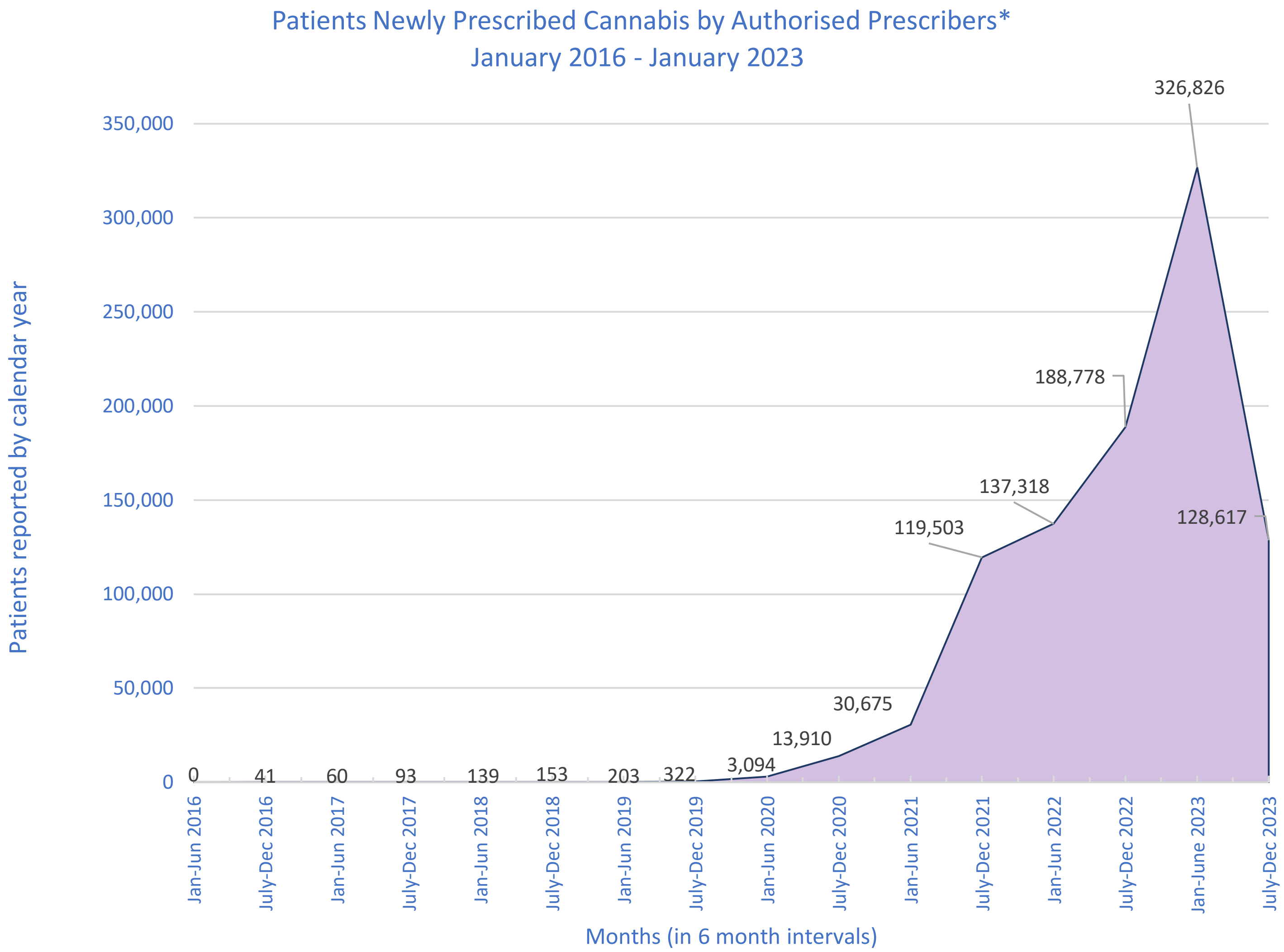

*Authorised Prescribers – Medical Practitioners approved by a HREC or endorsed by a specialist college to prescribe medicinal cannabis

Source: TGA. Medicinal Cannabis Authorised Prescriber Scheme data. Australian Government Therapeutic Goods Administration (TGA) 2023. Accessed September 07, 2023. https://www.tga.gov.au/products/unapproved-therapeutic-goods/medicinal-cannabis-hub/medicinal-cannabis-access-pathways-and-patient-access-data/medicinal-cannabis-authorised-prescriber-scheme-data

**Supplementary 2. PICO framework with keyword elements, inclusion, and exclusion criteria.**

| | | Keyword elements | Inclusion | Exclusion |
|---|---|---|---|---|
| **Population** | Countries allowing therapeutic usage of cannabis, prescribed or otherwise** | 1. Cannabis and cannabis-based products<br>2. Medical and therapeutic | Studies reporting on monitoring of cannabis use as a medicine for aforementioned indications. | Studies reporting on:<br>- Monitoring of recreational cannabis use.<br>- Synthetic Cannabinoid Receptor Agonists |
| **Intervention** | Systems for monitoring of side effects and adverse events | 3. Monitoring systems<br>4. Side effects and adverse events | Systems for monitoring of side effects and adverse events mentioned as a core component of the title or abstract. | Small scale surveys as a monitoring system<br>Studies relating to addictive potential of medicinal cannabis<br>Studies focusing on monitoring of adverse effects on social wellbeing (marriage, behaviour, academic achievement) |
| **Comparison** | No established monitoring system | N/A | N/A | N/A |
| **Outcome** | Integration of formalized monitoring systems into clinical practice (workflow) | 5. Pharmacovigilance | N/A | N/A |
| **Study design** | Peer-reviewed systematic reviews, scoping reviews, meta-analyses and narrative reviews, primary papers, grey literature | N/A | Papers published from Jan 2015 until June 2023. | Papers not published in English.<br>Animal studies<br>Poster presentations<br>Letters to the editor |

**Usage for the following indications: Chronic pain, cancer pain, epilepsy, neuropathic pain in MS, AIDS wasting syndrome, chemotherapy-induced nausea and vomiting, sleep disorders/insomnia, psychiatric disorders (PTSD, schizophrenia), inflammatory conditions such as rheumatoid arthritis (RA), Tourette's, irritable bowel syndrome (IBS), Parkinson's disease, dystonia, glaucoma, ADHD, autism.

**Supplementary 3. Categories of searches in search strategy.**

| **Category 1 - Cannabis, Medical, Monitoring Systems and Side Effects Keyword Elements** | | | |
|---|---|---|---|
| Cannabis search terms | Medical Keyword search terms | Monitoring system search terms | Side Effects and Adverse Events search terms |
| Cannabi* OR tetrahydrocannabinol OR THC OR delta9* OR nabiximol* OR dronabinol* or Cannabigerol* OR CBG OR CBN OR marijuana OR marihuana. | Therap* OR medic* OR prescrib* OR prescrip* OR treat* | Surveillance OR post marketing, safety, database, registry, registries, OR monitoring | Side effect* OR adverse effect* OR adverse outcome* OR adverse event* |
| **Category 2 - Cannabis, Medical, Monitoring Systems and Pharmacovigilance Keyword Elements** | | | |
| Cannabis search terms | Medical search terms | Monitoring search terms | Pharmacovigilance search terms |
| Cannabi* OR tetrahydrocannabinol OR THC OR delta9* OR nabiximol* OR dronabinol* or Cannabigerol* OR CBG OR CBN OR marijuana OR marihuana. | Therap* OR medic* OR prescrib* OR prescrip* OR treat* | Surveillance OR post marketing, safety, database, registry, registries, OR monitoring | Pharmacovigilance |

### Supplementary 4 Full electronic search strategy for OVID-Embase

#### Category 1 search

1. exp postmarketing surveillance/ or exp drug safety/
2. exp pharmacovigilance/ or exp drug surveillance program/
3. exp drug monitoring/
4. ("surveillance" or "post marketing" or "safety" or "database*" or "registry" or "registries" or "monitoring").ti,ab,kf.
5. 1 or 2 or 3 or 4
6. exp medical cannabis/ or exp cannabis derivative/ or exp "Cannabis sativa subsp. indica"/ or exp cannabis-induced psychosis/ or exp "Cannabis (genus)"/ or exp cannabis smoking/ or exp cannabis/ or exp "Cannabis sativa subsp. sativa"/ or exp "cannabis use"/
7. cannabinol/ or cannabinol derivative/ or exp cannabis/ or exp cannabinoid/ or "cannabis (genus)"/ or cannabis addiction/ or cannabis derivative/
8. ("cannabi*" or "tetrahydrocannabinol" or "THC" or "delta9*" or "nabiximol*" or "dronabinol*" or "cannabigerol*" or "CBG" or "CBN" or "marijuana" or "marihuana").ti,ab,kf.
9. 6 or 7 or 8
10. exp drug therapy/ or exp therapy/
11. ("therap*" or "medic*" or "prescrib*" or "prescrip*" or "treat*").ti,ab,kf.
12. 10 or 11
13. exp side effect/dm, si [Disease Management, Side Effect]
14. exp adverse event/ or exp adverse drug reaction/
15. ("side effect*" or "adverse effect*" or "adverse outcome*" or "adverse event*").ti,ab,kf.
16. 13 or 14 or 15
17. 5 and 9 and 12 and 16
18. limit 17 to yr="2015 - 2023"

#### Category 2 search

1. exp postmarketing surveillance/ or exp drug safety/
2. exp drug surveillance program/ or exp drug monitoring/
3. ("surveillance" or "post marketing" or "safety" or "database*" or "registry" or "registries" or "monitoring").ti,ab,kf.
4. 1 or 2 or 3
5. exp pharmacovigilance/
6. pharmacovigilance.ti,ab,kf.
7. 5 or 6
8. exp medical cannabis/ or exp cannabis derivative/ or exp "Cannabis sativa subsp. indica"/ or exp cannabis-induced psychosis/ or exp "Cannabis (genus)"/ or exp cannabis smoking/ or exp cannabis/ or exp "Cannabis sativa subsp. sativa"/ or exp "cannabis use"/
9. cannabinol/ or cannabinol derivative/ or exp cannabis/ or exp cannabinoid/ or "cannabis (genus)"/ or cannabis addiction/ or cannabis derivative/
10. ("cannabi*" or "tetrahydrocannabinol" or "THC" or "delta9*" or "nabiximol*" or "dronabinol*" or "cannabigerol*" or "CBG" or "CBN" or "marijuana" or "marihuana").ti,ab,kf.
11. 8 or 9 or 10
12. exp drug therapy/ or exp therapy/
13. ("therap*" or "medic*" or "prescrib*" or "prescrip*" or "treat*").ti,ab,kf.
14. 12 or 13
15. 4 and 7 and 11 and 14
16. limit 15 to yr="2015 - 2023"

### Supplementary 5. Monitoring systems included in primary papers

| Author/Date | Italian Phytovigilance Database | German Pain E-registry | Quebec Cannabis Registry | The Registry | Australian Emyria Clinical e-Registry | FAERS | UKMCR | ToxiC registry | Vigibase | SwissCanOn | Health Canada | Eudravigilance | TGA | AMK | AIFA | Project 21 |
|---|---|---|---|---|---|---|---|---|---|---|---|---|---|---|---|---|
| Sakal et al. 2021[79] | | | | | | | | | | | | | | | | ■ |
| Patti et al. 2016[77] | | | | | | | | | | | | | | | ■ | |
| Guevara et al. 2015[46] | | | | ■ | | | | | | | | | | | | |
| Obrien et al. 2023[78] | | | | | | | | | | | | | | | | ■ |
| Messina et al. 2017[76] | | | | | | | | | | | | | | | ■ | |
| Lynskey et al. 2022[68] | | | | | | | ■ | | | | | | | | | ■ |
| Ammendoliaetal.2023[49] | | | | | | | | | | | | ■ | | | | |
| Aprikianetal.2023[39] | | | ■ | | ■ | | | | | | ■ | | | | | |
| Cresciolietal.2020[28] | ■ | | | | | | | | | | | | | | | |
| Dalavayeetal.2023[61] | | | | | | | ■ | | | | | | | | | |
| Ergisietal.2022[63] | | | | | | | ■ | | | | | | | | | |
| Ergisietal.2023[52] | | | | | | | ■ | | | | | | | | | ■ |
| Erridgeetal.2022[58] | | | | | | | ■ | | | | | | | | | |
| Erridgeetal.2022[56] | | | | | | | ■ | | | | | | | | | |
| Erridgeetal.2021[50] | | | | | | | ■ | | | | | | | | | |
| Etgesetal.2016[45] | | | | ■ | | | | | | | | | | | | |
| vanHunseletal.2022[43] | | | | | | | | | ■ | | | | | | | |
| Freudewaldetal.2022[69] | | | | | | | | | | | | | | ■ | | |
| Jacketal.2022[47] | | | | | | | | | | | ■ | | | | | |
| Kawkaetal.2021[60] | | | | | | | ■ | | | | | | | | | |
| Rhyeeetal.2018[73] | | | | | | | | ■ | | | | | | | | |
| Loveetal.2022[72] | | | | | | | | ■ | | | | | | | | |
| Lunghietal.2022[35] | | | | | | ■ | | | | | | | | | | |
| Harrisetal.2022[66] | | | | | | | ■ | | | | | | | | | |

| |
|---|
| Mangooetal.2022[55] |
| Morideetal.2022[40] |
| Nicholasetal.2023[64] |
| Nimalanetal.2022[67] |
| Olssonetal.2023[51] |
| Pillaietal.2022[54] |
| Pochetetal.2022[44] |
| PrietoGonzalezetal.2021[33] |
| Pirachaetal.2017[37] |
| Rifkin-Zybutzetal.2023[57] |
| Holveyetal.2023[65] |
| Jacketal.2022[29] |
| Sharmaetal.2020[38] |
| Spyresetal.2020[71] |
| Spyresetal.2015[70] |
| Simonetal.2023[36] |
| Taitetal.2023[59] |
| Taitetal.2023[62] |
| TGA,2023.[74] |
| Trojanetal.2022[75] |
| Ueberalletal.2019[31] |
| Ueberalletal.2022[32] |
| Ueberalletal.2022[30] |
| Ueberalletal.2022[34] |
| Vickeryetal.2022[48] |
| Viganoetal.2020[41] |
| Viganoetal.2022[42] |
| Wangetal.2023[53] |

**Supplementary 6. Differences between databases and registries.**

| Characteristics | Database | Registry |
|---|---|---|
| **Purpose** | Often designed for multiple uses rather than only monitoring therapeutics | Designed with a specific purpose in mind, such as monitoring of effectiveness or safety |
| **Data structure** | Includes various data types, such as patient information, clinical trials data, and pharmacological details | Simpler and designed for a specific aim (i.e tracking patient outcomes over time for a specific condition or treatment) |
| **Flexibility** | May offer more flexibility, allowing complex queries that can filter several data types | Limited querying capabilities tailored to the specific data it collects |
| **Ownership** | Typically owned and created by the same institution | Owned by a variety of organizations but are often initiated by health agencies, research, or medical organisations |
| **Data Sources** | Multi-purpose, and can include both research and clinical data | Sources data directly from clinicians, patients, or specific clinical studies. |
| **Updates** | If not designed for real time monitoring, may not be regularly updated | Regularly updated for longitudinal monitoring |
| **Scope** | Wider in scope, not limited to a specific therapeutic area or patient population | Narrower in scope, focusing on a specific therapy or patient population. |

**Supplementary 7. Reporting hierarchy of side effects and adverse events associated with CBM usage**

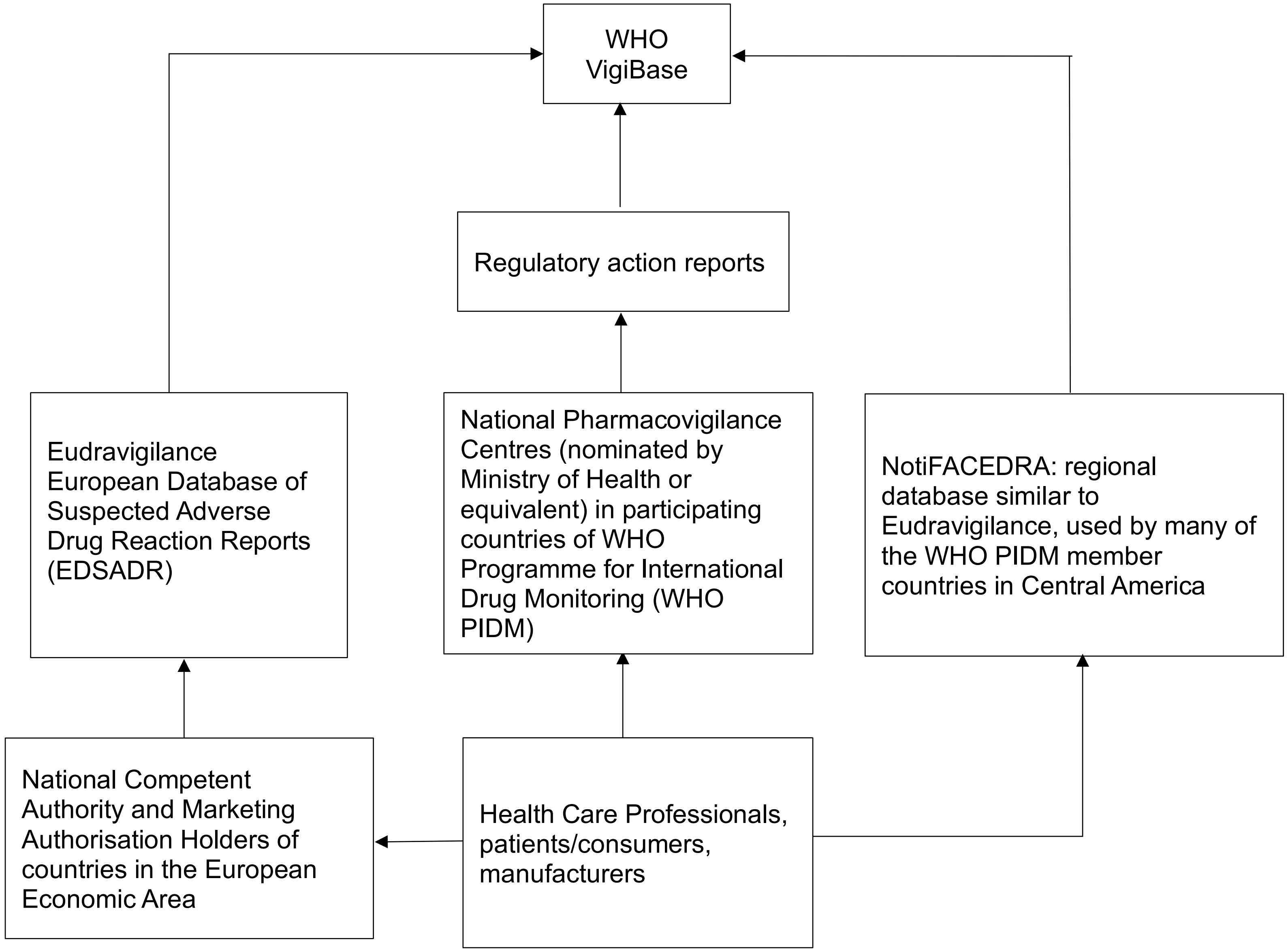
